# Artificial Intelligence Deployment of Conversational Support (AI-DOCS): A patient acceptability study

**DOI:** 10.1101/2025.07.19.25331814

**Authors:** Shrirajh Satheakeerthy, Andrew EC Booth, Liam Widdop, Shaun Evans, Brandon Stretton, Sarah Howson, Christina Gao, Rudy Goh, Toby Gilbert, Christopher Foster-McBride, Weng Onn Chan, Jamie W. Bellinge, Andrew Zannettino, John Maddison, Stephen Bacchi

## Abstract

This study evaluated an artificial intelligence (AI) system based on a large language model (LLM) in conducting complex patient assessments, comparing its acceptability to that of human basic physician trainees (BPTs) during a divisional clinical examination (DCE). Compared with BPTs, Artificial Intelligence Deployment of Conversational Support (AI-DOCS) scored similarly empathy, politeness, and comprehensiveness to human trainees. AI-DOCS also elicited important clinical information that was not divulged to human examiners. Further research is needed to explore the integration of AI-DOCS into clinical workflows.

## Introduction

Artificial intelligence (AI), particularly large language models (LLMs), has the potential to significantly enhance clinical workflows through conversational interactions with patients.^1^ Previous studies have shown that Artificial Intelligence Deployment of Conversational Support (AI-DOCS) can collate complex medical histories, synthesise information, and answer consultant questions at a level sufficient to pass the Royal Australasian College of Physicians (RACP) Divisional Clinical Examination (DCE), undertaken annually by basic physician trainees (BPTs).^2^ However, patient acceptability and experience of AI-DOCS remain unexplored.

Digital interventions must demonstrate measurable benefit prior to widespread implementation. For example, digital interventions have demonstrated benefits in reducing length of stay,^3^ preventing seven-day readmissions,^3^ improving weekend discharges,^4^ improving blood glucose level (BGL) control,^5^ identifying patients who will require dialysis,^6^ and reducing the inappropriate use of intravenous medications.^7^ LLMs present challenges in generating evidence for their implementation (including validation and reproducibility), in part due to the non-deterministic nature of proprietary frontier LLMs.^8^ Notwithstanding these hurdles, they have been shown to have significant utility in hospitals, such as in the automation of auditing.^9, 10^ Moving beyond clinician-facing tools to patient-facing AI systems raises distinct concerns, especially around acceptability and patient experience.

This study was conducted with the aim of evaluating the experience of patients who engage with AI-DOCS in an exam-style format and comparing this with the human physician trainees undertaking the same task.

## Methods

The study was conducted with approval from the Central Adelaide Local Health Network Human Research Ethics Committee (reference number 20824) and individual patient consent.

This single-site study was performed at the Lyell McEwin Hospital, a tertiary teaching hospital and accredited training provider for physicians undertaking ‘BPT’, which includes consultant-led trial examinations. These trial examinations emulate the format of the RACP DCE, with approximately half of the examination consisting of ‘long cases’. In a long case a trainee has 60 minutes with a complex patient to take a comprehensive history, perform a rigorous examination and then formulate a presentation. Candidates are usually provided with a medication list but otherwise rely upon their clinical skills and reasoning for the evaluation.

The study was undertaken at the final trial examination prior to the annual RACP DCE. The patient volunteers were first evaluated by AI-DOCS. The same patients were then evaluated by trial examination candidates.

AI-DOCS was delivered via a laptop or tablet using *Anthropic*’s Claude Sonnet 4 model as the LLM, which was provided with an overview of the process of a long case. AI-DOCS is unable to perform a physical examination and therefore undertook the cases similar to the format of a “phone long case”. This format was employed by the RACP during the COVID pandemic and involved a candidate undertaking a history over the phone, while physical examination findings (and the baseline medication list and vital signs) were provided in writing. The physical examination findings provided to AI-DOCS were those described by a clinician familiar with the patient. To compensate for the time typically allocated to physical examination, AI-DOCS had a reduced time of 40 minutes with the patient. Communication with the AI-DOCS language model was autonomous and no intervention was provided by study investigators to assist the conversation.

Participating patients completed a survey involving Likert-type responses and open-ended questions relating to their experience with both AI-DOCS and the human physician examinee. The questions included the topics of politeness, comprehensiveness and empathy (scale of 0 to 5). A score of how likely a patient would be to recommend the doctor or AI to a friend was also collected on a 0 to 10 scale.

Human examiners (either at an advanced trainee, qualified physician, or a consultant level) evaluated BPTs based on the RACP DCE format. It is a 25-minute period comprising a verbal presentation and question answering (with usually approximately half the time dedicated to each component). Examiners would also evaluate AI-DOCS presenting the case, and answering questions verbally, with a subsequent examiner free-text description of feedback on its presentation. Candidate-level results are reported with descriptive statistics, with inclusion of all numerical results in the manuscript to enable future reanalyses.

## Results

Six of seven eligible patients consented to participate and were evaluated by AI-DOCS. One patient was subsequently not seen by human physician trainees due to logistical reasons, meaning five patients were seen by both physician trainees and AI-DOCS. Additionally, difficulty recruiting examiners meant that only four of the cases evaluated by AI-DOCS were presented to human examiners. There were four female patients (4/5, 80%). The medical conditions of this cohort included amyloidosis, heart transplant, renal transplant, pancreas transplant, Parkinson’s disease, and various chronic complications of immunosuppression and infections.

In terms of empathy, the BPTs scored 4.50 (IQR 4.00–5.00), while the LLM scored 5.00 (IQR 5.00–5.00) (see Table 1). The BPTs received a politeness score of 5.00 (IQR 5.00– 5.00), compared with 5.00 (IQR 5.00–5.00) for the LLM. For comprehensiveness, the BPTs scored 5.00 (IQR 4.25–5.00) versus 5.00 (IQR 5.00–5.00) for the LLM. Ratings of how likely patients would be to recommend the doctor or AI to a friend were 10.00 (IQR 9.25–10.00) for the BPTs and 10.00 (IQR 5.00–10.00) for the LLM.

**Table 1.** Results from patient surveys regarding LLM and human trainee performance.

| Patient | Candidate | Politeness<br>(maximum 5) | Comprehensiveness<br>(maximum 5) | Empathy<br>(maximum 5) | Likelihood of<br>recommending<br>(maximum 10) | Comments |
| --- | --- | --- | --- | --- | --- | --- |
| A | LLM | 5 | 5 | 5 | 10 | A Truly amazing experience |
| A | Human | 5 | 5 | 5 | 10 | - |
| A | Human | 5 | 5 | 5 | 10 | - |
| B | Human | 5 | 5 | 5 | 10 | - |
| B | LLM | 5 | 5 | 5 | 10 | - |
| B | Human | 5 | 5 | 5 | 10 | - |
| C | Human | 5 | 4 | 4 | 8 | - |
| C | LLM | 5 | 5 | 5 | 10 | - |
| C | Human | 4 | 5 | 4 | 8 | - |
| D | LLM | 4 | 4 | 5 | 5 | It depends on your attitude towards computers and new technology like AI. |
| D | Human | 5 | 4 | 5 | 10 | used a few medical terms that I was not familiar with, had to clarify |
| D | Human | 5 | 4 | 4 | 9 | - |
| E | LLM | 5 | 5 | 5 | 5 | not sure that AI is something I want going forward. |
| E | Human | 5 | 5 | 4 | 10 |  |
| E | Human | 5 | 5 | 4 | 10 |  |

Comments from patients suggested that AI-DOCS would be more approachable with a human name. Some patients said their willingness to recommend AI-DOCS to a friend, would depend on the individual’s attitude to technology, rather than the AI itself. Positive comments were included about the empathy of AI-DOCS.

Difficulty recruiting human examiners resulted in feedback on only four of the cases. Narrative feedback from examiners was generally positive with statements including “The AI has done an amazing job with the structure, presentation and flow of [redacted]’s history – if I were sitting the exam again, I’d be listening to this presentation a few more times to try and emulate the style of presentation and descriptive terms used” and “Dangerously exciting (will I be replaced by AI) to see what AI can do with complex medical information.” Critical feedback of AI-DOCS in the presentation phase related to the inclusion of unnecessary content on the mechanism of action of medications. Additionally, examiners fed back that AI-DOCS elicited information from patients that they had not elicited. In particular, this additional information related to mental health concerns, with one patient disclosing a history of suicidality to AI-DOCS that had not been disclosed to the human examiners. In accordance with ethics and legal requirements, and additionally with the patient’s consent, this information was provided to the patient’s primary healthcare provider, who was aware of and managing the issue.

## Discussion

This small pilot study has shown that AI-DOCS, an LLM-based platform, is rated by patients as having a communication ability during complex clinical histories comparable to that of senior physician trainees. AI-DOCS also elicited information not obtained by human examiners. These findings support the potential utility of the approach and further investigation.

While the trial DCE provided a useful setting to evaluate AI-DOCS at this stage, it should be noted that the examination setting is different from real clinical practice. Trainees may be nervous and are particularly conscious of time in examination settings. While time-constraints exist in clinical practice, exam conditions may result in less empathetic communication from trainees.

Patients who volunteer for long cases are “practised” patients, in that they usually are selected for being able to describe their medical history effectively and in English. Patients with cognitive impairment, dysarthria or dysphasia, or difficulties communicating in English are often excluded from participation in the DCE. Conversely, patients are selected for long cases due to their extraordinary medical and social complexity.

It would be academically interesting to evaluate the clinical performance of AI-DOCS as compared to physician trainees in this setting, as adjudicated by human examiners. However, in this examination environment, it was impractical to recruit sufficient numbers of candidates and experienced examiners to make a statistical comparison useful. Previous studies have already shown that AI can perform at a standard sufficient to pass RACP long cases.^2^

Further research is warranted to evaluate technologies like AI-DOCS in clinical settings. Avenues for exploration include smartphone applications or phone calls to facilitate patient reviews, particularly in the outpatient setting.

## Conclusions

AI-DOCS, an LLM-based conversational platform, interviewed complex medical patients in the format of an RACP DCE and had similar scores to human BPTs on comprehensiveness, politeness, and empathy. AI-DOCS also elicited important information from patients that was not disclosed to human examiners. Further research examining how such approaches could augment existing clinical workflows is required.

## Data Availability

Data sharing are limited by patient confidentiality and ethical approvals.

